# Mistletoe as a Therapeutic Option in Cancer Patients: a scoping review protocol

**DOI:** 10.1101/2023.07.29.23293280

**Authors:** Milena Mendes Cordeiro, Felipe Aguiar Pupo Seabra Malta, Daniela Caetano Gonçalves

**Author notes:** M.M.C. and F.A.P.S.M. contributed equally.

## Abstract

**Objective:** This scoping review aims to identify, map and provide an overview of the studies investigating the therapeutic use of Viscum Album L. (mistletoe) in cancer patients. By identifying pivotal patterns and research gaps in the existing literature on Viscum therapy, this review intends to offer valuable insights and supportive information to better inform future research efforts in this field.

**Introduction:** Cancer exerts a significant impact on global mortality, posing numerous challenges to public policies. The disease progression and the conventional treatment itself presents multifaceted health adversities to subjects. In response to these complexities, researchers have been exploring non-conventional therapeutic approaches, including phytotherapy. Mistletoe, a widely-used herbal medicine in oncology, is currently undergoing investigation for its potential adjuvant roles in both curative and palliative cancer care.

**Inclusion criteria:** We will consider studies that include individuals with cancer, irrespective of participant’s age or cancer type/stage, and who were subjected/exposed to interventions based on or incorporating Viscum album L. The review will consider clinical, quasi-experimental, and analytical observational studies, with no limitations on the time of intervention/exposure, treatment stage or geographic region.

**Methods:** This review will follow the methodological guidance of the JBI Manual for Evidence Synthesis for scoping reviews. The searches will be conducted in the following databases: MEDLINE (via PubMed), CENTRAL (via Cochrane Library), Embase (via Elsevier), Scopus, Web of Science Core Collection, CINAHL (via EBSCO), LILACS and SciELO. The search strategy will be adapted to the specificities of each of these databases, using thesaurus terms and synonyms related to the intervention (mistletoe) and the population (cancer), without restrictions on language or date of publication. The search for unpublished studies will be done in Google Scholar, preprint servers (medRxiv and bioRxiv) and trial registry platforms (Clinical Trials and International Clinical Trials Registry Platform), along grey literature in OpenGrey. Initially, eligibility criteria will be screened by two independent reviewers, assessing article titles and abstracts. Following finishing the full reading and inclusion determination, data extraction will be carried out by two independent reviewers. The results will be presented in narrative form and through tables and graphs.

## INTRODUCTION

Cancer is a disease that stands as one of the primary causes of death worldwide, posing a challenge to increasing life expectancy. Global estimates suggested that approximately 19.3 million new cancer cases emerged in 2020, with around 10 million deaths reported. The most prevalent cancer types include breast, prostate, colorectal and lung, in addition to melanoma (Sung et al., 2021). Cancer development is influenced by major risk factors, which can be categorized as either exogenous, such as smoking, alcohol consumption, ultraviolet radiation, and diet, or endogenous factors, including aging and genetic susceptibility (Lewandowska et al., 2019).

The process of carcinogenesis involves multiple stages and is initially characterized by accumulation of changes caused by mutations and/or errors in chromosome segregation during mitosis in the genome, mainly in genes that play a role in maintaining cellular balance, such as those involved in cell death and proliferation. Cancer cells typically exhibit adaptive advantage to escape host defense mechanisms, undergo uncontrolled proliferation, regulate angiogenesis, invade adjacent tissues, and metastasize (Fares et al., 2020); (N Kontomanolis et al., 2021).

Apart from its cellular, metabolic, and physiological repercussions, cancer has negative effects on patients’ health, resulting in various symptoms, including pain, dyspnea, fatigue, nausea, sleep disturbances, psychological alterations, weight and appetite loss (Walsh & Rybicki, 2006). To deal with the disease, the main treatment options are surgical interventions, chemotherapy, radiotherapy, and immunotherapy. The selection and the success of these modalities rely on numerous variables, depending on the tumor type/stage and its behavior, as well as the patient’s general health status and the timing of the intervention (Debela et al., 2021). Nevertheless, the treatment itself gives rise to various side effects, negatively impacting the autonomy and quality of life of the subjects (Cheng et al., 2017).

Fatigue, defined as a distressing, persistent, subjective sense of physical, emotional, and/or cognitive tiredness or exhaustion that is not proportional to recent activity and interferes with usual functioning, is one of these major symptoms (Berger et al., 2015). Additionally, other effects, including pain, cognitive impairment, anorexia, hot flashes, peripheral neuropathy, lymphedema, hair loss, insomnia, and social isolation, also have an impact on the personal and social aspects of the cancer patient (Jacobs & Shulman, 2017).

In this context, complementary and alternative therapies emerge as options to be used in conjunction with conventional treatment. Non-conventional therapeutic approaches, such as naturopathy, Traditional Chinese Medicine (TCM) and phytotherapy (Kuo et al., 2018), aim to address not only curative intentions but also to alleviate disease symptoms and treatment side effects, promoting well-being and enhancing quality of life (Hiam-Galvez et al., 2021); (Greenlee et al., 2017).

Phytotherapy, the therapeutic use of plants or plant extracts, usually produced from minimal industrial processing, has gained prominence due to its cultural tradition and wide accessibility to the population (Lopes et al., 2017); (Tilburt & Kaptchuk, 2008). In cancer, plants like Echinacea, Ginseng, Allium sativum L., and Viscum album L. are among the most commonly used. Specifically, mistletoe (Viscum album L.) is widely employed in cancer treatment, with a focus on curative and palliative treatment (Nazaruk & Orlikowski, 2016).

Viscum album L. is a hemiparasitic plant that belongs to the family Santalaceae (Felenda et al., 2019). Viscum is typically administered via subcutaneous injection, and its utilization varies depending on the processing method, such as aqueous extraction, pressing, and fermentation of the aqueous extract. Also, the therapy can be employed in isolated forms containing specific bioactive components, such as lectins, as for example ML-1 (mistletoe lectin-1), or viscotoxins, such as VT-A (viscotoxin A) (Felenda et al., 2019; Ostermann et al., 2019, 2020).

In cancer research, mistletoe’s therapeutic potential has been explored across multiple tumor types, such as breast, lung, and pancreatic (Matthes et al., 2020). This investigation has covered various contexts, including adjuvant treatment, focusing on its antitumor actions and potential to enhance survival rates, as well as palliative care, to alleviate adverse treatment and disease symptoms, such as fatigue, aiming to improve overall quality of life (Horneber et al., 2008).

A preliminary search of MEDLINE, the Cochrane Database of Systematic Reviews and JBI Evidence Synthesis was conducted and no current or underway systematic reviews or scoping reviews on the topic were identified.

## REVIEW QUESTION

1. What are the main characteristics of the participants included (e.g. cancer type and stage, treatment phase, gender, and age)?
2. What are the key characteristics of Viscum as an intervention/exposure? (e.g. isoform, timing, dose, frequency, and route of administration).
3. What outcomes are commonly reported and what assessment tools have been applied to measure them?
4. What are the methodological designs of studies utilized in the construction of evidence (e.g. randomized controlled trials, pre-test/post-test, prospective cohort, etc.)?
5. What is the profile of the authors conducting research on the topic (e.g. nationality, affiliated institutions, potential conflicts of interest, funding sources, etc.)?

## ELIGIBILITY CRITERIA

### Participants

This review aims to be inclusive, considering cancer patients of all ages, genders, and demographic backgrounds. It will cover the different cancer types and stages, investigating various clinical contexts, from the time of diagnosis to the period of remission. Notably, the review will comprehensively consider both curative and palliative treatment purposes, assessing diverse modalities such as drug therapy, radiotherapy, chemotherapy, immunotherapy, surgery, and more.

### Concept

Interventions utilizing Viscum album L. or its active components. The interventions may be administered alone or in combination with other conventional and non-conventional therapies, irrespective of their timing, cycle or route of administration.

### Context

This study review aims to provide a comprehensive global analysis, covering the topic across multiple geographical regions. It will encompass diverse settings, including hospitals, hospices, tertiary institutes, home care, among others. There will be no restrictions on the year of publication or the language used in the studies.

### Types of Sources

This scoping review will consider both experimental and quasi-experimental study designs including randomized controlled trials, before and after studies and interrupted time-series studies. In addition, analytical observational studies including prospective and retrospective cohort studies, case-control studies and analytical cross-sectional studies will be considered for inclusion.

## METHODS

The proposed scoping review will be conducted in accordance with the JBI methodology for scoping reviews and will be reported following the guidelines of the Preferred Reporting Items for Systematic Reviews and MetaAnalyses extension for Scoping Reviews (PRISMA-ScR) (Tricco et al., 2018).

### Search strategy

The search strategy will aim to locate both published and unpublished studies. An initial limited search of MEDLINE (PubMed) and Embase was undertaken to identify articles on the topic. The text words related to the intervention (mistletoe) and the population (cancer) contained in the titles and abstracts of relevant articles, and the index terms used to describe the articles were used to develop a full search strategy for CINAHL, MEDLINE, LILACS, SciELO, Embase, Scopus, CENTRAL e Web of Science. The search strategy, including all identified keywords and index terms (MeSH and Emtree), will be adapted for each included database and/or information source (Table 1). Also, screening for additional studies will be done in Google Scholar, grey literature databases (OpenGrey), trial registry platforms (Clinical Trials and International Clinical Trials Registry Platform) and preprint servers (bioRxiv and medRxiv). Authors of papers will not be contacted to request missing or additional data. Studies published in any language and at any date will be included.

**Table 1.**
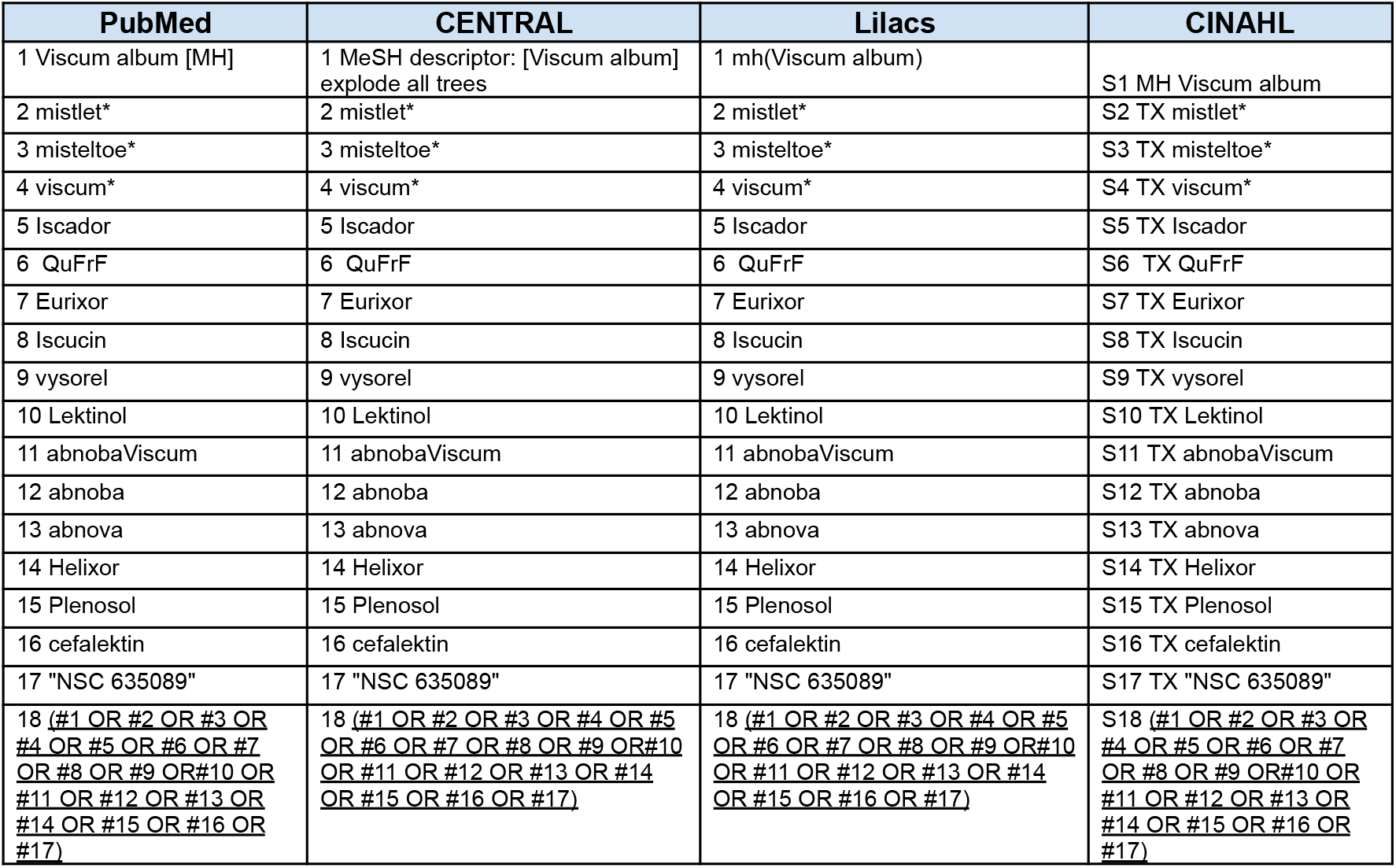
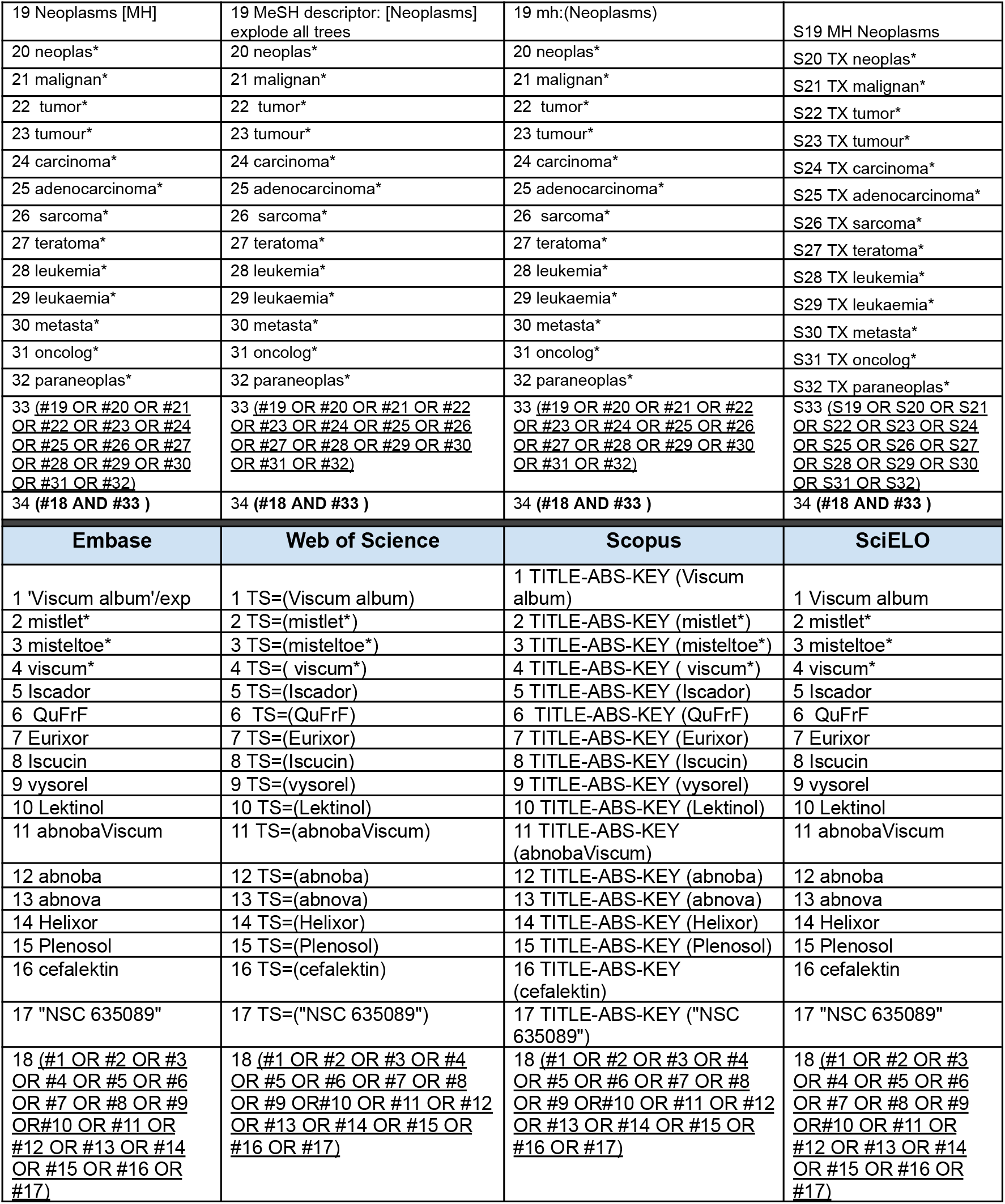
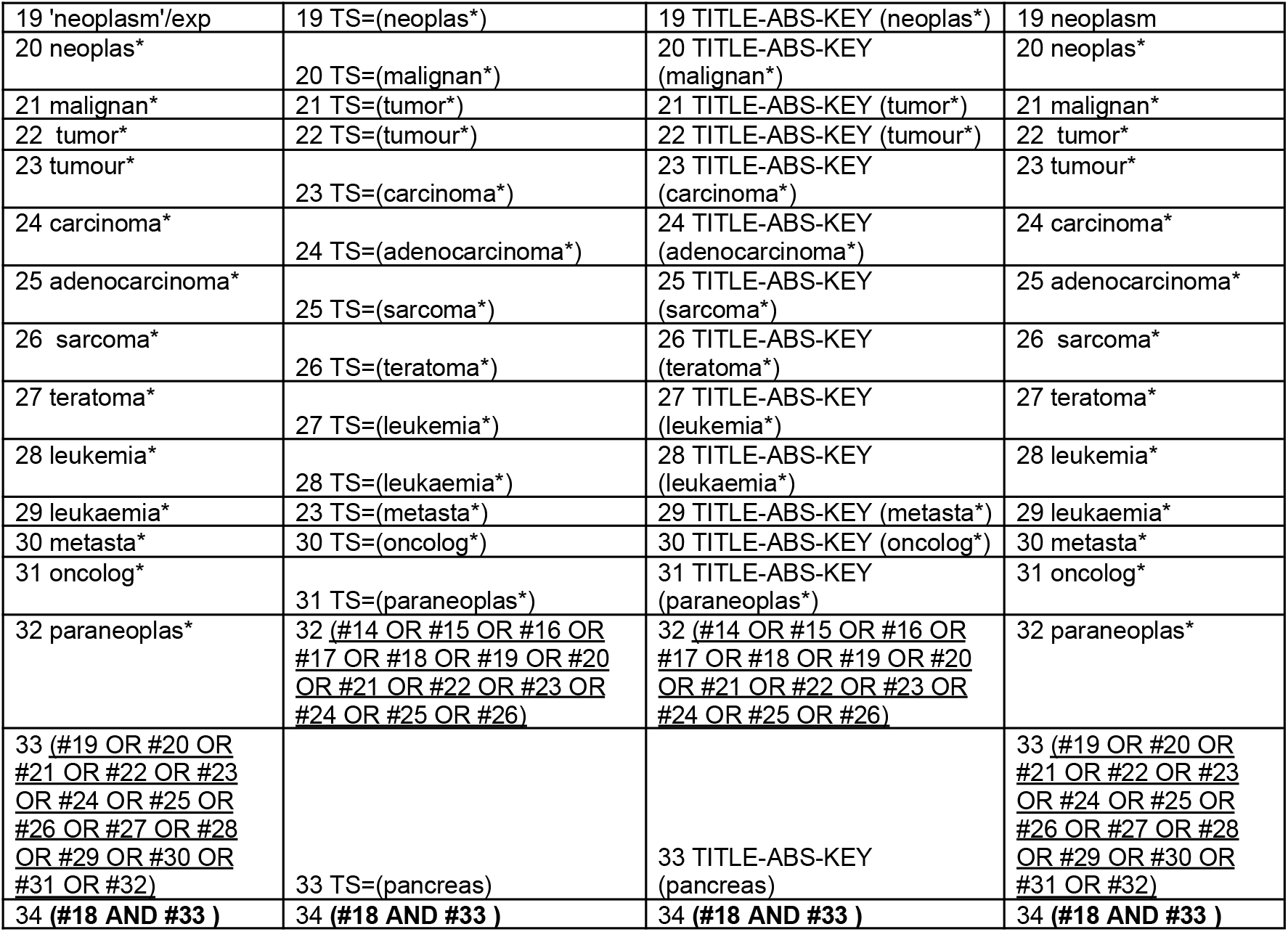
Search strategy.

### Study/Source of Evidence selection

All identified records will be uploaded into Mendeley (Sungur & Seyhan, 2013) and duplicates removed. After deduplication, records will be inserted into Rayyan (Ouzzani et al., 2016). Following a re-check and removal of additional duplicated articles, titles and abstracts will then be screened by two independent reviewers for assessment against the inclusion criteria for the review (Mendes M. and Malta F.). Potentially relevant sources will be retrieved in full. The full text of selected citations will be assessed in detail against the inclusion criteria by two researchers (Mendes M. and Malta F.). Reasons for exclusion of sources of evidence at full text that do not meet the inclusion criteria will be recorded and reported in the scoping review. In case of persistent disagreement, a third reviewer (Caetano. D) will be contacted to mediate the discussion or determine the final decision. The results of the search and the study inclusion process will be reported in full in the final scoping review and presented using the PRISMA-ScR flow diagram (Tricco et al., 2018).

### Data Extraction

Data will be extracted from records included in the scoping review by two reviewers (Mendes M. and Malta F.) using a data extraction tool developed by the reviewers. The data extracted will include specific details about the participants (age, sex, cancer type/stage, etc.), concept (viscum isoform, dose, frequency, etc.), context (study country, cancer treatment context, physical location, etc.), study methods (design, follow-up, outcomes evaluated, etc.) and key findings relevant to the review question/s (journal, year of publication, funding, pre-register, etc.). A draft extraction form is provided (Table 2). The draft data extraction tool will be modified and revised as necessary during the process of extracting data from each included evidence source. Modifications will be detailed in the scoping review. Any disagreements that arise between the reviewers will be resolved through discussion, or by consulting the third reviewer (Caetano G.). Due to the high number of records, the authors of the studies will not be contacted to request missing or additional data, where required.

**Table 2.** Data extraction instrument.

| Domain | Variable |
| --- | --- |
| **Publication Characteristics** | Year of publication [integer] |
|  | Journal Name [string] |
|  | Journal Nationality [string] |
|  | Authors' Name [list] |
|  | Authors' Nationality [list] |
|  | International Collaboration [binary — Yes, No] |
| **Study Characteristics** | Design [string] |
|  | Primary Outcome [string] |
|  | Period of Realization [date] |
|  | Number of Participating Centers [integer] |
|  | Country of Participating Centers [list] |
|  | Level of Care of Participating Centers [Primary, Secondary, Tertiary, Quaternary] |
|  | Sample Size [integer — number of participants enrolled] |
|  | Attrition Rate [double — proportion of attrition rate] |
|  | Funding [list] |
|  | Private Funding [binary — Yes, No] |
|  | Conflict of Interest Reported [binary — Yes, No] |
|  | Ethical Statements [list] |
|  | Preregistration [binary — Yes, No] |
| **Population** | Gender [double — proportion of women] |
|  | Age, range [string] |
|  | Age, mean [double] |
|  | Cancer Type [string] |
|  | Cancer Stage [string] |
|  | Treatment Objective [binary — curative, palliative] |
|  | Treatment Context [string — e.g. preoperative, post surgery, during chemotherapy] |
|  | Treatment Context Details [string — e.g. type of surgery/chemotherapy] |
| **Intervention** | Sample size [integer] |
|  | Total Duration [double — number of weeks] |
|  | Dose [string] |
|  | Frequency [string] |
|  | Route of Administration [string] |
|  | Intervention Details [string] |
|  | Presence of Multitherapy [binary — Yes, No] |
|  | Multitherapy type [string] |
| Control | Sample size [integer] |
|  | Type of control [Placebo, No-treatment, Active, External, Multiple, Dose-response] |
|  | Description [string] |
| Outcomes | Reported Outcomes [list] |
|  | Method of Assessment [list] |

### Data Analysis and Presentation

The extracted data will align with the review’s objective and questions outlined in this protocol. The included studies will be summarized in tabular form, providing general characteristics such as publication details, study design, population, intervention, and outcomes assessed. Moreover, to address the study objectives and questions, capturing characteristics of the conceptual categories aforementioned in this protocol, we will present summary statistics (absolute and relative frequencies), tables, charts (heatmap matrices, bar and line graphs), and figures (infographics, maps). Additionally, a qualitative synthesis will be performed to describe the key findings, patterns, and gaps identified in the existing literature.

## Data Availability

All data produced in the present work are contained in the manuscript.

## Acknowledgements

Nassib Bezerra Bueno, for valuable advice regarding the development of this manuscript.

## LIST OF TABLES

- Table 1. Search strategy
- Table 2. Data extraction instrument

